# Biventricular Pressure-Volume Loop Analysis Predicts Outcomes After Double Switch Operation For Congenitally Corrected Transposition Of The Great Arteries with Intact Ventricular Septum

**DOI:** 10.1101/2025.03.17.25324148

**Authors:** Nikhil Thatte, Peter E. Hammer, Gerald Marx, Rebecca Beroukhim, Kimberlee Gauvreau, Ryan Callahan, Ashwin Prakash, Sitaram Emani, David Hoganson, Pedro del Nido, Sunil J. Ghelani

## Abstract

**Background:** Assessing left ventricular (LV) preparedness in congenitally-corrected transposition of the great arteries/intact ventricular septum (ccTGA/IVS) prior to the double switch operation (DSO) remains challenging. Subpulmonary LV Pressure-Volume Area (PVA) - a comprehensive metric of ventricular workload - when compared to systemic right ventricular (RV) PVA as a benchmark, may be a good index of adequacy.

**Aims:** Determine (1) if LV PVA can be estimated from simple catheterization and imaging parameters, using conductance-catheter derived PVA as reference, and (2) if LV:RV ePVA ratio predicts outcomes after DSO.

**Methods:** Subpulmonary LV PVA was measured using conductance catheters and compared to estimated PVA (ePVA) calculated with simple catheterization and volumetric variables. Then, in a retrospective cohort, LV:RV ePVA ratio and other clinical variables were evaluated as predictors for a composite adverse outcome of ≥moderate LV dysfunction, transplant, or death post-DSO.

**Results:** ePVA yielded high agreement and low bias compared to measured PVA by conductance catheter (n=20). In the retrospective cohort, 6/42 patients (14%) experienced the outcome. Low LV:RV ePVA and pressure ratios were the only significant predictors, while LV mass and mass-to-volume ratio were not. Amongst 8 patients with borderline pressure ratios, ePVA ratio was an excellent discriminator – five with ePVA ratio <0.67 had adverse outcome, whereas three with ePVA ratio ≥0.67 did not.

**Conclusions:** Estimation of subpulmonary LV PVA using simple imaging and catheterization data was reliable compared to gold standard techniques. LV:RV ePVA ratio ≥0.67 was a strong and novel predictor of LV preparedness for DSO in patients with ccTGA/IVS.

**Central Message:** Biventricular pressure-volume area (PVA), a surrogate of myocardial V̇O2, can be estimated in ccTGA patients. LV:RV PVA ratio >0.67 is a novel marker of LV preparedness for double switch operation.

**Perspective Statement:** The current approach for determining LV preparedness prior to double switch operation (DSO) is inadequate with a 15% rate of post-DSO LV dysfunction. Pressure-Volume Area (PVA) is a comprehensive yet concise metric incorporating both pressure and volume work. Use of LV:RV PVA ratio may improve patient selection and therefore outcomes after DSO in patients with ccTGA and intact ventricular septum.

## 1 Introduction

The optimal management strategy for congenitally corrected transposition of the great arteries (ccTGA) with intact or nearly intact ventricular septum (IVS) remains a subject of debate in the congenital heart disease community. In cases without associated anomalies, a conservative approach—leaving the morphologic right ventricle (RV) in the systemic (subaortic) position—has been the traditional choice. However, this approach is associated with a high incidence of late systemic RV dysfunction.^1,2^

An alternative strategy is anatomic repair through a double switch operation (DSO), which combines both atrial and arterial switch procedures to restore the morphologic left ventricle (LV) as the systemic ventricle and the RV as the subpulmonary ventricle. While the technical outcomes of DSO are excellent, approximately 15% of patients experience LV failure following the procedure.^2–5^

In the absence of a ventricular septal defect or LV outflow tract obstruction, deconditioning of the subpulmonary LV occurs relatively rapidly within the first months of life. Therefore, in order to prepare the LV for DSO, a pulmonary artery band (PAB) is placed to impose additional afterload on the LV.^6–8^ Multimodal assessment, including echocardiography, cardiac magnetic resonance imaging (MRI), and cardiac catheterization, uses standard metrics such as LV to RV pressure ratio, LV mass, and LV mass-to-volume ratio to evaluate LV preparedness for DSO.^2^

Pressure-Volume Area (PVA) by pressure-volume (PV) loop analysis is a measure of total ventricular contractile energy expenditure (supplemental figure 1). It correlates closely with myocardial oxygen consumption (V̇O_2_).^9,10^ PVA incorporates both the pressure and volume change components of ventricular work in a single metric. For patients undergoing DSO, the systemic RV’s PVA could serve as an internal control to estimate the contractile energy demands on the LV in the post-DSO physiology. This concept led us to hypothesize that an inadequate LV:RV PVA ratio is predictive of adverse outcomes after DSO. However, direct measurement of LV PVA requires the use of micromanometer-tipped conductance catheters, limiting its clinical use in centers without this technological expertise as well as the ability to test this variable as a predictor of outcomes in a retrospective cohort where such data were not already obtained. We further hypothesized that LV PVA could be estimated accurately using standard imaging and catheterization data.

Thus, we sought to determine if (1) LV PVA can be estimated (ePVA) from simple catheterization and imaging parameters, using conductance-catheter derived PVA as reference, and (2) if LV:RV ePVA ratio predicts outcomes after DSO.

## 2 Methods

### 2.1 Study Design

This was a single-center retrospective cohort study conducted in two phases (Figure 1).

**Figure 1.**
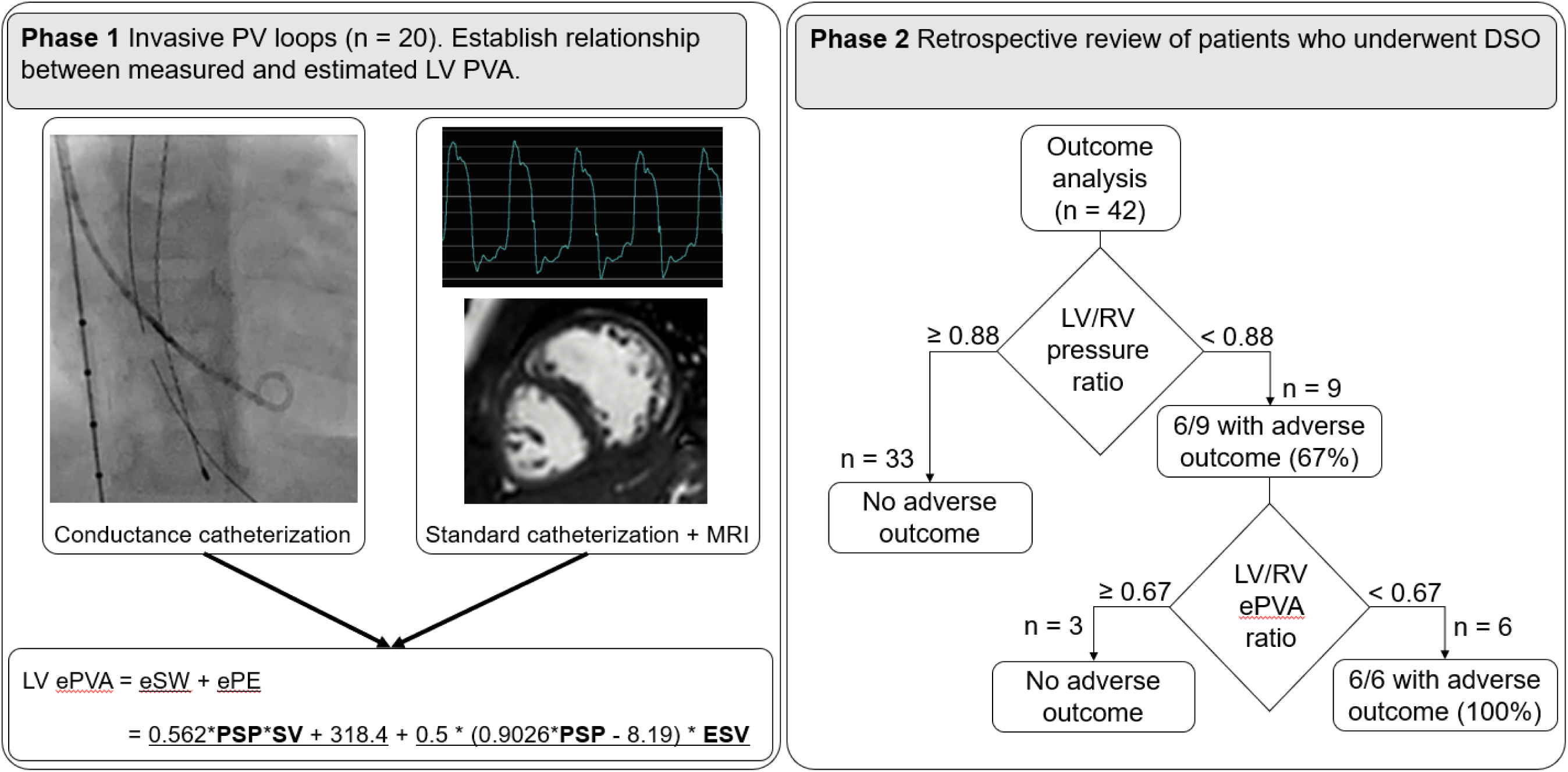
Central illustration. Legend: This study evaluated the feasibility of computing pressure-volume area (PVA) using standard catheterization and MRI parameters, and then utilizing a novel LV:RV ePVA ratio to determine preparedness of subpulmonary LVs for double switch operation in patients with congenitally corrected transposition of the great arteries with intact ventricular septum. Abbreviations: MRI, magnetic resonance imaging; PV, pressure volume; LV, left ventricle; ePVA, estimated pressure-volume area; eSW, estimated stroke work; ePE, estimated potential energy; PSP, peak systolic pressure; SV, stroke volume; ESV, end-systolic volume; RV, right ventricle; DSO, double switch operation

Phase 1: Patients with ccTGA/IVS (pre-DSO) undergoing cardiac catheterization as part of a routine pre-DSO workup between 2021-2023 were included for PV-loop analysis in the subpulmonary LV using a conductance catheter (CD Leycom ®, Netherlands). Linear regression was used to correlate directly measured PVA with estimations using parameters measured easily during standard catheterization and volumetric imaging. Patients with large LVs leading to measured stroke work >5000 mmHg.mL were excluded as these outliers would skew the regression equations for the majority of the remainder of the cohort of predominantly young children.

Phase 2: All patients with ccTGA/IVS who underwent DSO at our institution between 1990 and 2023 were included for a retrospective outcome analysis. Patients who did not have both a pre-DSO catheterization and a pre-DSO biventricular volumetric imaging study (cardiac MRI, cine CT, or 3D echocardiogram) to allow computation of PVA were excluded. Patients with pre-existing moderate or worse LV systolic dysfunction pre-DSO were excluded. The surgical strategy and techniques for PA banding and DSO at our center have been previously published.^3^ Clinical, imaging, catheterization, and surgical data were collected from the electronic medical record. LV ePVA was calculated using the equation derived from phase 1. Follow-up data to determine outcomes were obtained from the electronic medical record.

This study was approved by the Boston Children’s Hospital Institutional Review Board. The requirement for individual patient consent was waived.

### 2.2 PV Loops Acquisition

Under general endotracheal anesthesia, a micromanometer-tipped conductance catheter (CD Leycom ®, Netherlands), was inserted into the subpulmonary LV via a long sheath in the right internal jugular vein. Volume calibration was performed using LV volumes taken from cardiac MRI, cine CT, or 3D echocardiogram performed prior to the catheterization. Loops were recorded with breath holding at end-expiration. Data were acquired for at least 10 steady state beats and averaged. Recordings with ectopic beats were excluded. Loops were analyzed using proprietary analysis software (ConductNT, CD Leycom ®, Netherlands).

#### 2.3.1 LV energetics

The total mechanical energy expenditure of a cardiac beat – the Pressure-Volume Area (PVA) – is the sum of external stroke work (SW) which is the area bounded within a PV loop, and the potential energy (PE) of contraction which is the triangular area on the origin side of the PV loop bounded by the end-systolic PV point and V_0_ (the volume-axis intercept of the end-systolic pressure volume relationship (ESPVR) (supplemental figure 1).^9^ End-systole was defined as the point on the PV loop where the ratio of pressure to volume was highest. LV SW (mmHg.mL) was directly measured (measured SW, mSW) from the recorded PV loops; LV PE (mmHg.mL) was measured (mPE), assuming V_0_ = 0, as mPE = 0.5 * LV end-systolic pressure (ESP) * LV end-systolic volume (ESV). Measured PVA (mPVA) was calculated as mSW + mPE.

#### 2.3.2 Systemic RV PVA estimation

For the systemic RV, SW was calculated as aortic mean arterial pressure * stroke volume using previously established methods for SW estimation in normal systemic ventricles.^11^ RV PE was calculated as 0.5 * RV ESV * RV ESP where RV ESP was calculated as 0.9 * aortic peak systolic pressure (assuming no significant aortic stenosis).^12^ RV PVA was then calculated as RV PVA = RV SW + RV PE.

#### 2.3.3 PVA Estimation Methodology

The goal of this analysis was to estimate LV PVA (ePVA) using data easily obtained by catheterization (LV pressures) and imaging (LV volumes), using data recorded by conductance catheter (mSW + mPE = mPVA) as reference. Stroke work, Potential Energy, and PVA estimations are described in the Appendix. ePVA was then calculated as eSW + ePE and compared with mPVA.

### 2.4 Outcomes

The primary outcome was a composite adverse outcome of moderate or worse LV dysfunction, heart transplant, or death after DSO. Risk factors for the composite outcome studied included demographics such as age at first PA band, time from first PA band to DSO, age at DSO, pre-DSO imaging parameters including indexed LV mass and mass-to-volume ratio, presence of advanced 2^nd^ degree or complete heart block, pacemaker status, pre-DSO LV:RV pressure ratio, LV:RV ePVA ratio, surgical variables including cardiopulmonary bypass time and cross-clamp time.

### 2.5 Statistical Analysis

Phase 1: The estimations for SW, PE, and PVA were evaluated by Pearson correlation and Bland-Altman analyses, using directly measured SW, PE, and PVA from conductance catheter-derived PV loops as reference.

Phase 2: Continuous data are presented as mean ± standard deviation or median [IQR], and categorical variables as number (percent). Fisher’s exact test was used to compare categorical variables, the Wilcoxon rank sum test for ordinal variables and for continuous variables that were not normally distributed, and Student’s two-sample t test for normally distributed continuous variables. Univariate time-to-event analyses were performed using Cox proportional hazards regression to determine variables significantly associated with the outcome. Discrimination was quantified using the c index. A multivariable model including the two strongest candidate predictor variables was not informative as these two variables (LV:RV pressure ratio and LV:RV ePVA ratio) were strongly correlated with one another. Bivariable models containing each of these ratios in combination with each other candidate predictor were studied. Analyses were performed using SAS Version 9.4 (SAS Institute, Inc) and GraphPad Prism version 10 (GraphPad Prism Software, La Jolla, California).

## 3 Results

### 3.1 LV PVA measurement and estimation

20 invasive PV loop studies from 18 unique patients with a median age of 2.4 [1.7, 5.4] years and median weight of 12.2 [11.1, 17.5] kg were recorded (Table 1). The equations derived from the linear regression methods along with their Pearson r correlation coefficients and Bland-Altman analyses are shown in supplemental figure 4. This yielded the following formula for estimation of LV PVA:

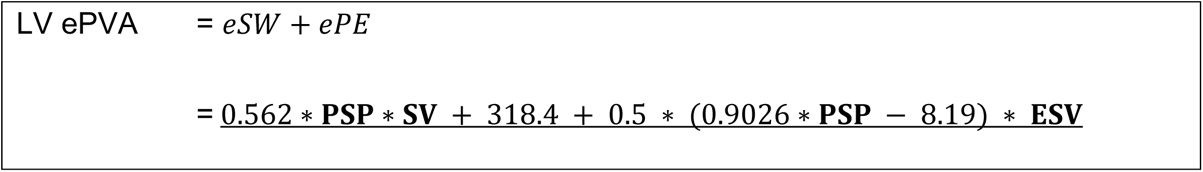

**Table 1.**
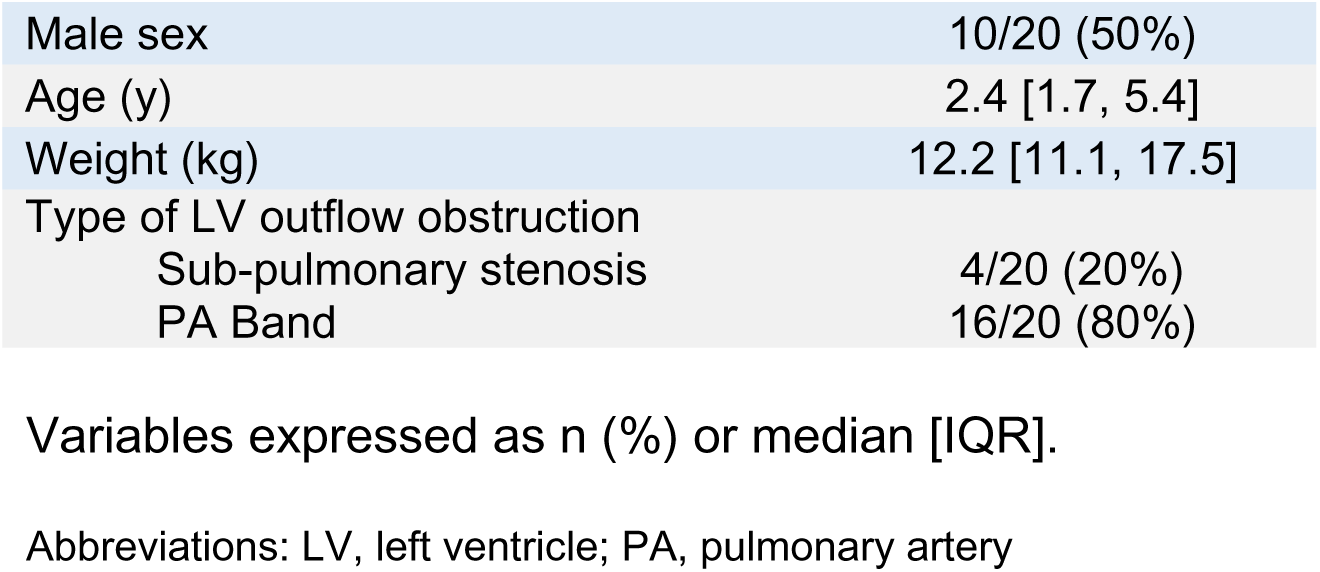
Baseline characteristics of patients in whom micromanometer-tipped conductance catheter studies were performed (n=20 studies in 18 unique patients).

|  |  |
| --- | --- |
| Male sex | 10/20 (50%) |
| Age (y) | 2.4 [1.7, 5.4] |
| Weight (kg) | 12.2 [11.1, 17.5] |
| Type of LV outflow obstruction |  |
| Sub-pulmonary stenosis | 4/20 (20%) |
| PA Band | 16/20 (80%) |

Variables expressed as n (%) or median [IQR].
Abbreviations: LV, left ventricle; PA, pulmonary artery

This equation uses only three simple LV metrics – peak systolic pressure (PSP), stroke volume (SV) and end-systolic volume (ESV) to calculate the LV ePVA.

This yielded excellent correlation and agreement with mPVA, R^2^ = 0.975, p<0.0001, Pearson r = 0.99, mean bias by Bland-Altman analysis 0.54 ± 11% (supplemental figure 4).

Using the above equation, the biventricular ePVA ratio is calculated as follows:

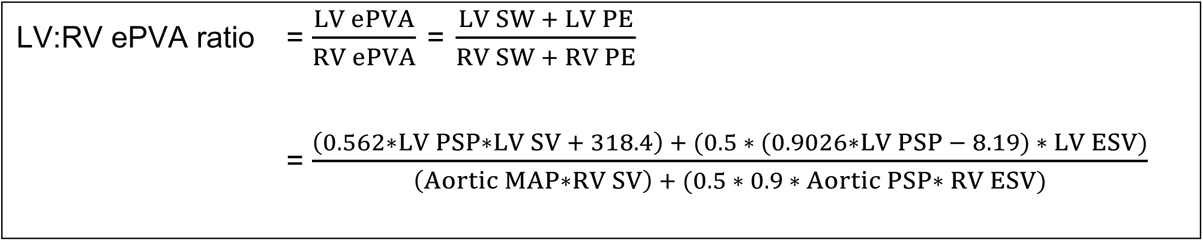

As intracardiac pressures vary throughout a catheterization case, as best as possible the aortic pressures (mean arterial pressure, MAP and peak systolic pressure, PSP) are chosen to be those which are simultaneous measurements with the LV PSP measurement to allow the most accurate comparison (similar to calculation of an LV:RV pressure ratio).

### 3.2 Outcomes of DSO and its predictors

Baseline characteristics of the patients are summarized in Table 2. 6/42 patients (14%) reached the composite outcome at a median follow up of 1.65 [0.12, 4.58] years. There were no significant differences between the groups in demographic factors, native anatomy, age at DSO, age at PA band, time from first PA band to DSO, whether or not an ASD was created at time of PA band placement, conduction and pacemaker status, and surgical parameters such as cardiopulmonary bypass time and cross-clamp time. Traditional volumetric and catheterization predictors of preparedness are summarized in Tables 3 and 4, along with LV:RV ePVA ratio. LV:RV ePVA ratio (0.57 [0.49, 0.61] vs 0.90 [0.73, 1.1], p<0.001) and LV:RV pressure ratio (0.80 [0.77, 0.87] vs 1.13 [1.01, 1.23], p<0.001) differed between the outcome groups. Results of univariate time-to-event analyses are summarized in Table 5. LV:RV ePVA ratio was the most significant determinant of outcome (HR 39 per 1 unit decrease, p = 0.004, C Index 0.94), while LV:RV pressure ratio was the only other significant predictor (HR 24 per 1 unit decrease, p = 0.005, C Index 0.88). No differences were present in the purely volumetric measures such as indexed LV mass and LV mass-to-volume ratio. On bivariable modeling, no other candidate risk factor was statistically significant in combination with either of these two parameters.

**Table 2.**
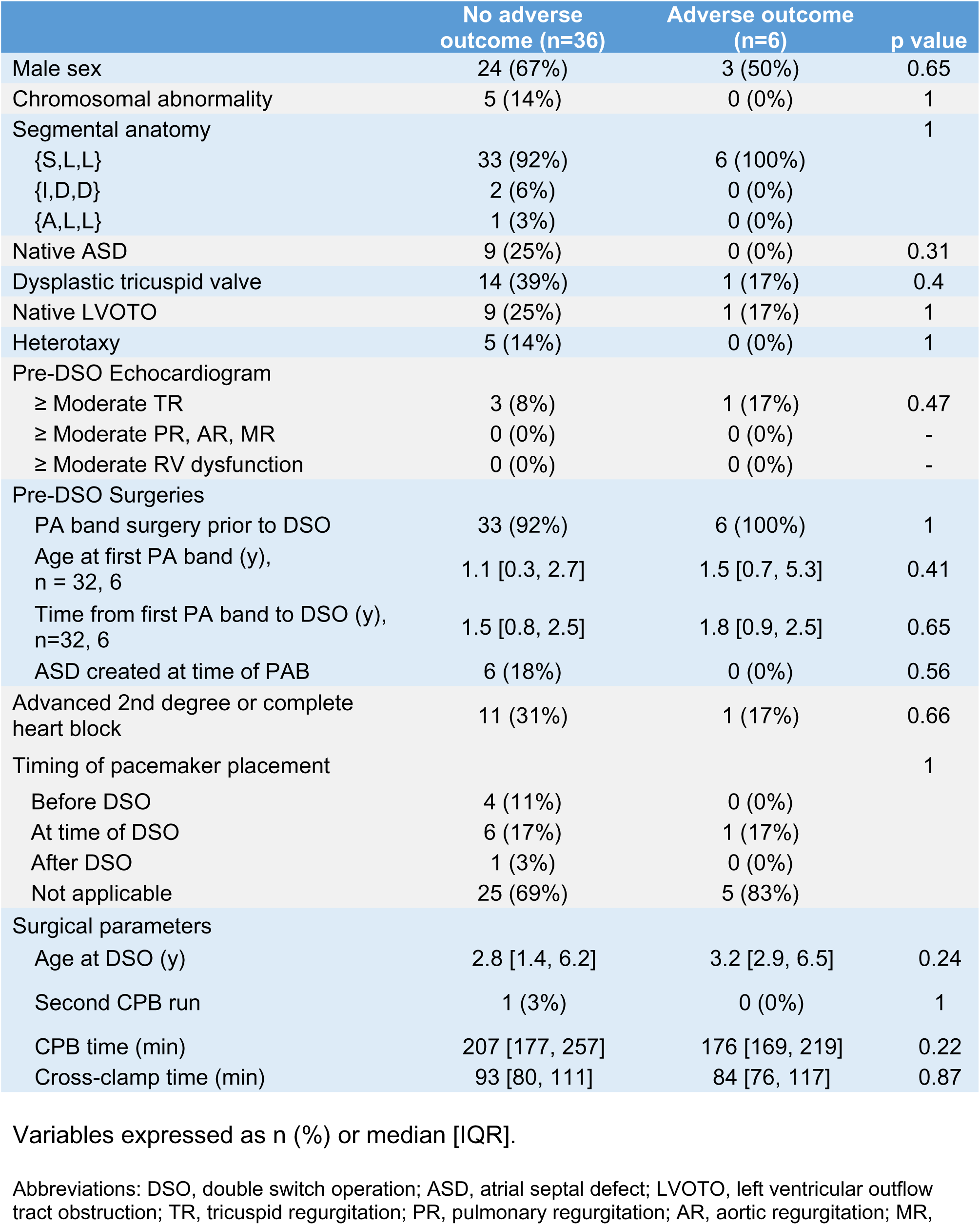
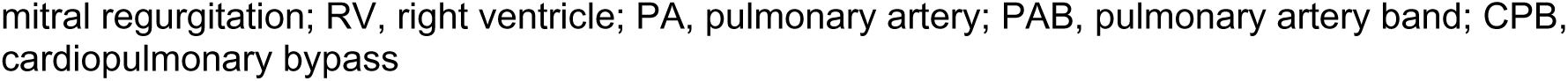
Baseline characteristics of patients prior to DSO (n=42).

|  | No adverse outcome (n=36) | Adverse outcome (n=6) | p value |
| --- | --- | --- | --- |
| Male sex | 24 (67%) | 3 (50%) | 0.65 |
| Chromosomal abnormality | 5 (14%) | 0 (0%) | 1 |
| Segmental anatomy |  |  | 1 |
| {S,L,L} | 33 (92%) | 6 (100%) |  |
| {I,D,D} | 2 (6%) | 0 (0%) |  |
| {A,L,L} | 1 (3%) | 0 (0%) |  |
| Native ASD | 9 (25%) | 0 (0%) | 0.31 |
| Dysplastic tricuspid valve | 14 (39%) | 1 (17%) | 0.4 |
| Native LVOTO | 9 (25%) | 1 (17%) | 1 |
| Heterotaxy | 5 (14%) | 0 (0%) | 1 |
| Pre-DSO Echocardiogram |  |  |  |
| ≥ Moderate TR | 3 (8%) | 1 (17%) | 0.47 |
| ≥ Moderate PR, AR, MR | 0 (0%) | 0 (0%) | - |
| ≥ Moderate RV dysfunction | 0 (0%) | 0 (0%) | - |
| Pre-DSO Surgeries |  |  |  |
| PA band surgery prior to DSO | 33 (92%) | 6 (100%) | 1 |
| Age at first PA band (y), n = 32, 6 | 1.1 [0.3, 2.7] | 1.5 [0.7, 5.3] | 0.41 |
| Time from first PA band to DSO (y), n=32, 6 | 1.5 [0.8, 2.5] | 1.8 [0.9, 2.5] | 0.65 |
| ASD created at time of PAB | 6 (18%) | 0 (0%) | 0.56 |
| Advanced 2nd degree or complete heart block | 11 (31%) | 1 (17%) | 0.66 |
| Timing of pacemaker placement |  |  | 1 |
| Before DSO | 4 (11%) | 0 (0%) |  |
| At time of DSO | 6 (17%) | 1 (17%) |  |
| After DSO | 1 (3%) | 0 (0%) |  |
| Not applicable | 25 (69%) | 5 (83%) |  |
| Surgical parameters |  |  |  |
| Age at DSO (y) | 2.8 [1.4, 6.2] | 3.2 [2.9, 6.5] | 0.24 |
| Second CPB run | 1 (3%) | 0 (0%) | 1 |
| CPB time (min) | 207 [177, 257] | 176 [169, 219] | 0.22 |
| Cross-clamp time (min) | 93 [80, 111] | 84 [76, 117] | 0.87 |

Variables expressed as n (%) or median [IQR].
Abbreviations: DSO, double switch operation; ASD, atrial septal defect; LVOTO, left ventricular outflow tract obstruction; TR, tricuspid regurgitation; PR, pulmonary regurgitation; AR, aortic regurgitation; MR,
mitral regurgitation; RV, right ventricle; PA, pulmonary artery; PAB, pulmonary artery band; CPB, cardiopulmonary bypass

**Table 3.** Volumetric imaging metrics.

|  | No adverse outcome<br>(n=36) | Adverse outcome (n=6) | p value |
| --- | --- | --- | --- |
| Indexed LV EDV, ml/m<sup>2</sup> | 69 [55, 88] | 63 [59, 77] | 0.36 |
| LV EF, % | 63 [55, 68] | 63 [61, 64] | 0.99 |
| Indexed LV mass, gm/m<sup>2</sup> | 38 [32, 55] | 34 [30, 42] | 0.26 |
| LV mass/volume ratio | 0.56 [0.45, 0.72] | 0.50 [0.48, 0.61] | 0.63 |

Variables expressed as median [IQR].
Abbreviations: LV, left ventricle; EDV, end-diastolic volume; EF, ejection fraction

**Table 4.** Traditional catheterization and PV-loop metrics.

|  | No adverse outcome<br>(n=36) | Adverse outcome (n=6) | p value |
| --- | --- | --- | --- |
| Cardiac index, ml/min/m<sup>2</sup> | 3.5 [3.2, 4.1] | 3.8 [3.5, 4.2] | 0.27 |
| LV EDP <10 mmHg | 13 (36%) | 3 (50%) | 0.66 |
| **LV:RV pressure ratio** | **1.13 [1.01, 1.23]** | **0.80 [0.77, 0.87]** | **<0.001** |
| **LV:RV ePVA ratio** | **0.90 [0.73, 1.10]** | **0.57 [0.49, 0.61]** | **<0.001** |

Variables expressed as n (%) or median [IQR].
Abbreviations: LV, left ventricle; EDP, end-diastolic pressure; RV, right ventricle; ePVA, Estimated pressure-volume area

**Table 5.**
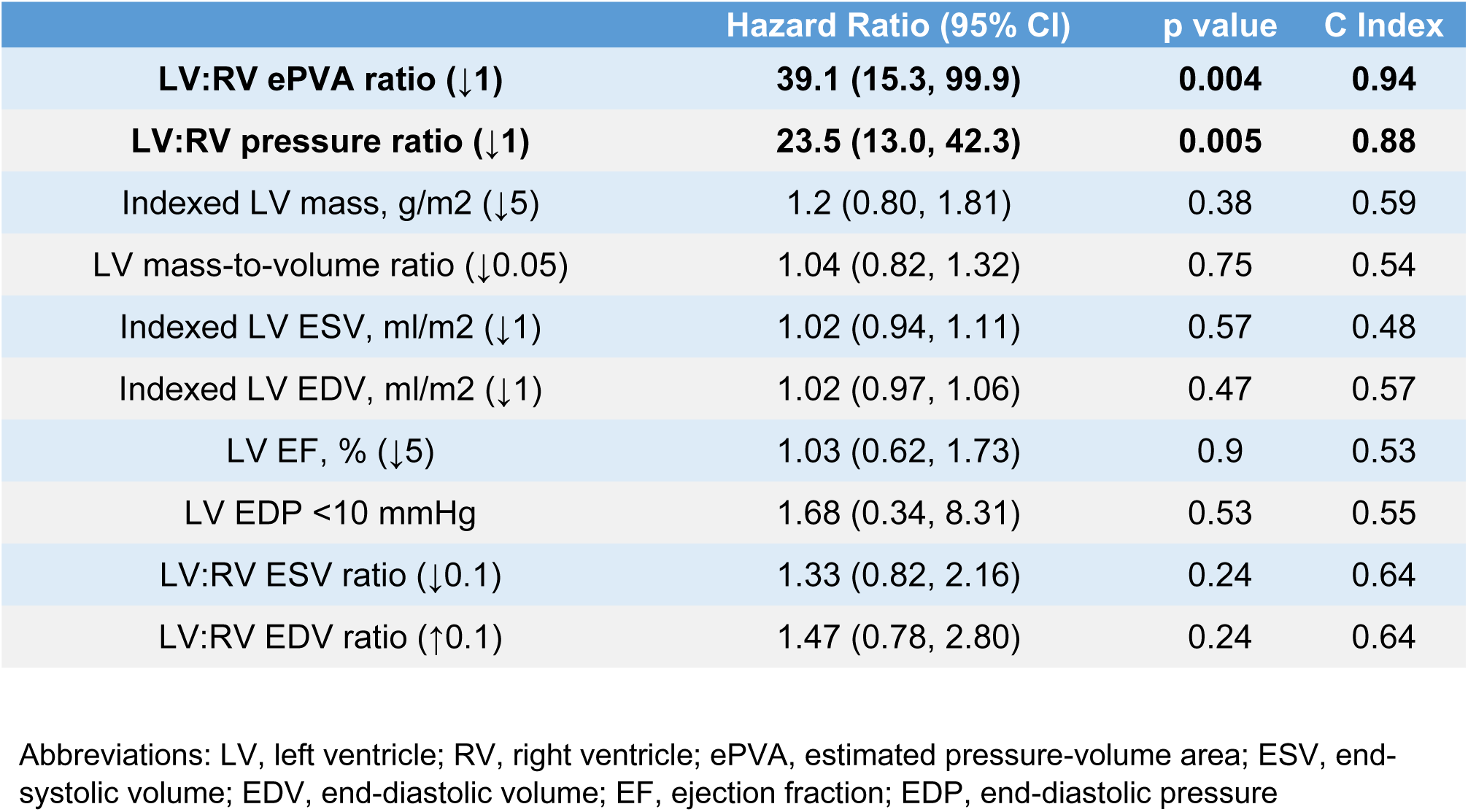
Predictors of composite adverse outcome by univariate time-to-event analysis (N=42).

An LV:RV ePVA ratio of 0.67 and LV:RV pressure ratio of 0.88 were found to be excellent binary thresholds (Figure 2); no patient with an LV:RV ePVA ratio ≥ 0.67 or pressure ratio ≥ 0.88 had an adverse outcome.

**Figure 2.**
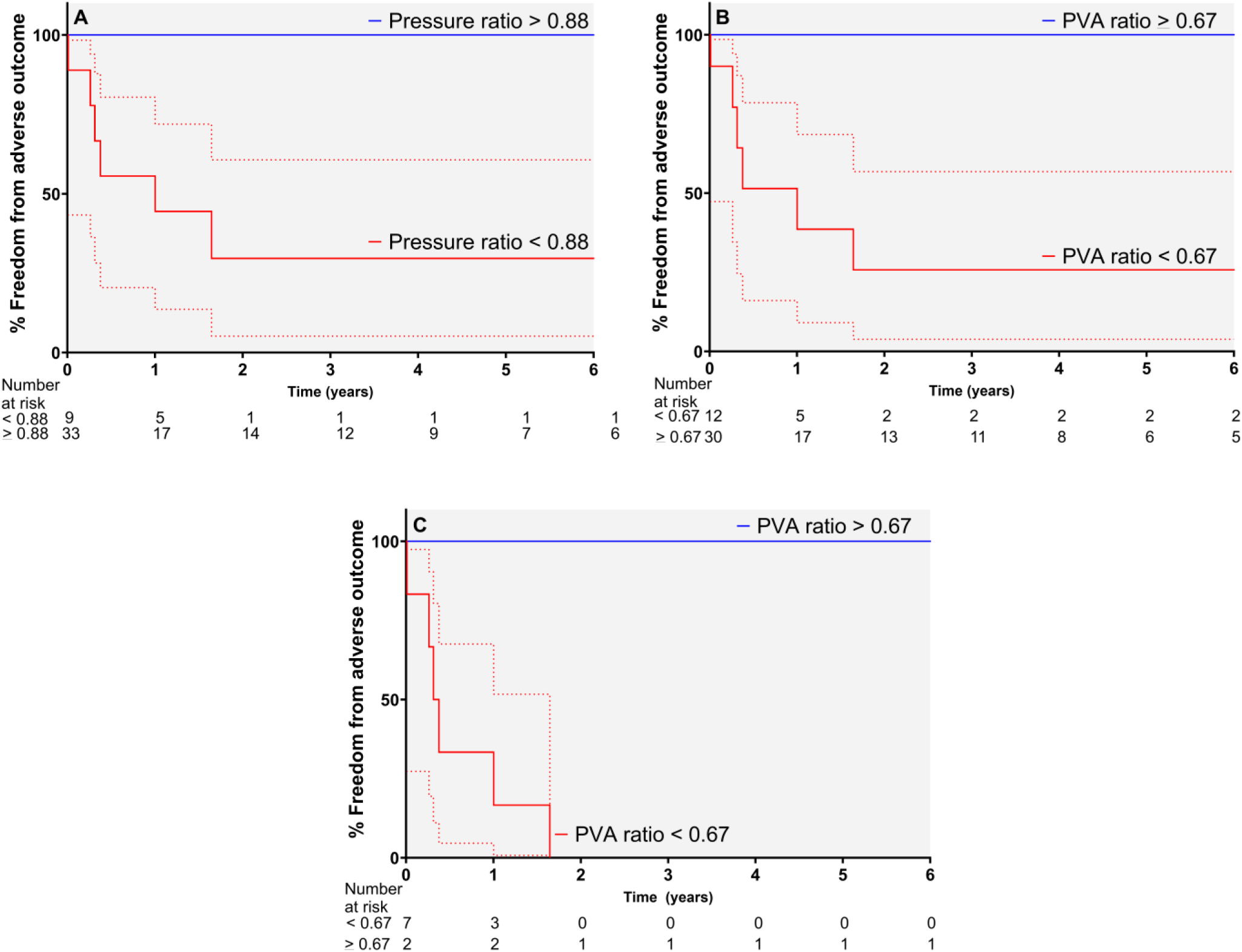
Freedom from composite adverse outcome in patients stratified by LV:RV pressure ratio (Panel A) and LV:RV ePVA ratio (Panel B). Panel C shows the subset of patients who had LV/RV pressure ratio <0.88, stratified by ePVA ratio. Abbreviations: LV, left ventricle; RV, right ventricle; PVA, pressure-volume area

Amongst those with a borderline LV:RV pressure ratio (n=8, 0.77-0.88), where clinical decision-making regarding LV preparedness may be challenging, the LV:RV ePVA ratio was an excellent discriminating parameter. In this subgroup, 3/8 (37%) patients who had adequate LV:RV ePVA ratios of 0.68, 0.73, and 1.03 did well, whereas 5/8 (63%) patients who had low LV:RV ePVA ratios of 0.44 to 0.60 had adverse outcomes (Figure 3). Four illustrative patients with biventricular PV loop analysis are depicted in Figure 4.

**Figure 3.**
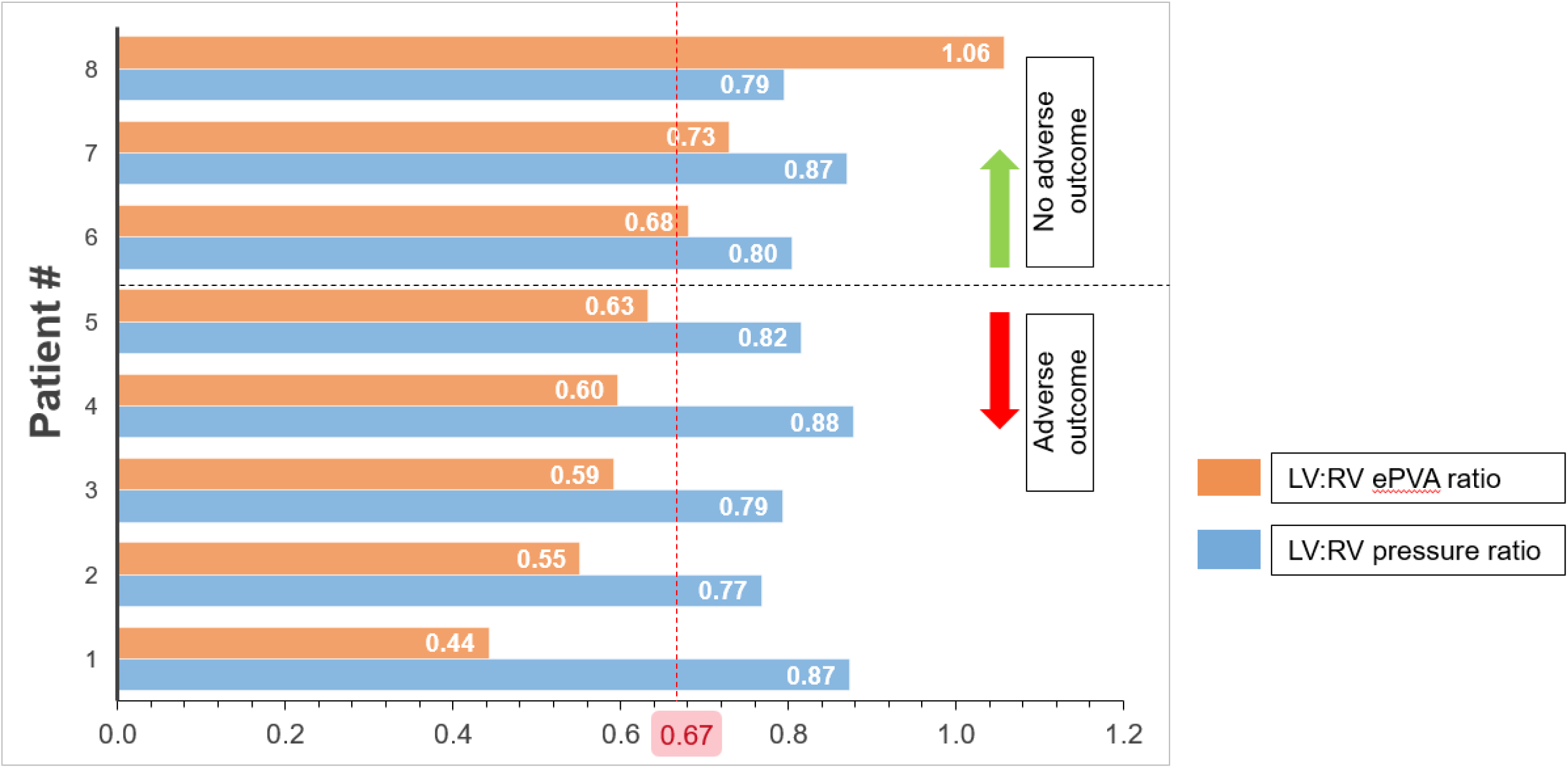
Bar graph depicting LV:RV pressure ratio (blue) and estimated Pressure-Volume Area ratio (orange) for 8 patients with challenging clinical decision making. Legend: Amongst those patients with borderline LV:RV pressure ratio of 0.77-0.88 (n=8), ePVA ratio was an excellent discriminating parameter. Patients with adequate LV:RV ePVA ratios (patient #6-8) had good outcomes, whereas those with low ePVA ratios (patient #1-5) had adverse outcomes. Abbreviations: LV, left ventricle; RV, right ventricle; ePVA, estimated pressure-volume area ratio

**Figure 4.**
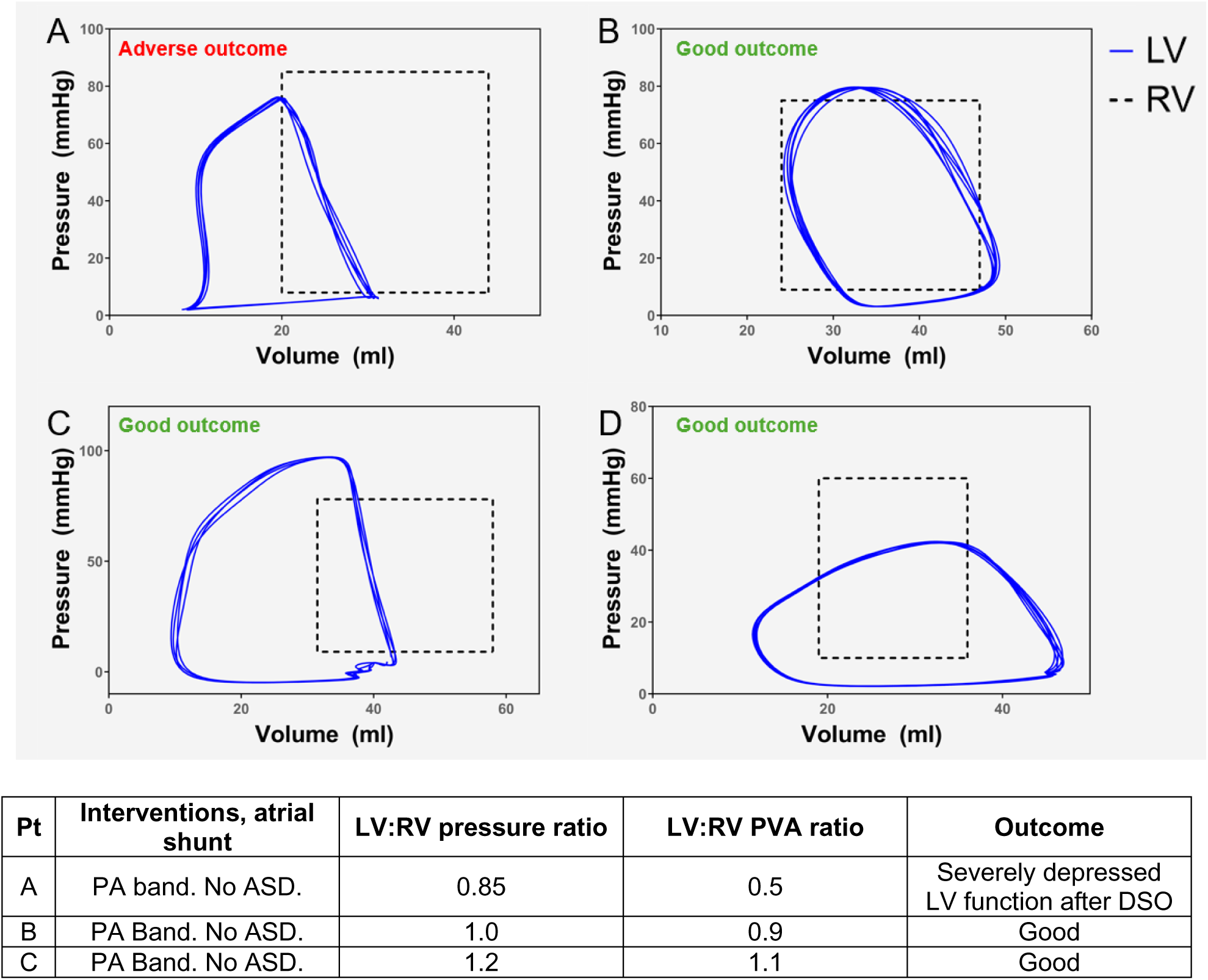
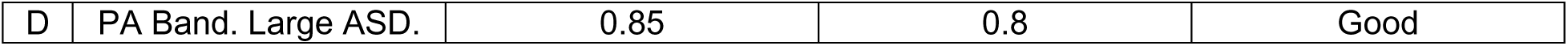
Case examples illustrating the utility of a PV-loop based visualization of biventricular work, described by the LV:RV PVA ratio, in predicting DSO outcomes. Legend: Subpulmonary LV loops (blue) were directly recorded using conductance catheters. RV loops (dashed black) were estimated as an idealized rectangle using catheterization pressures and MRI volumes. Patient A had a PA band with LV pressure at 85% of systemic pressure. Biventricular stroke volumes were equal (no shunts), and the LV:RV PVA ratio was low at 0.5. Despite a borderline adequate LV:RV pressure ratio and favorable LV EF, mass, and mass-to-volume ratios, the patient had an adverse outcome after DSO with severe LV dysfunction. Patients B, C, and D, with adequate LV:RV PVA ratios, demonstrated good outcomes. Patient D had borderline LV:RV pressure ratio of 0.85 similar to Patient A, but had a larger size LV due to a large ASD that led to greater stroke work and a better LV:RV PVA ratio of 0.8. In contrast, Patients B and C exhibited systemic or suprasystemic LV pressures and had no atrial level shunts. Patient B shows mild LV dilation (LV volumes match RV volumes) while Patient C had suprasystemic LV pressure without LV dilation (smaller LV end-diastolic volume compared to their RV). Abbreviations: PV, pressure-volume; PVA, pressure-volume area; MRI, magnetic resonance imaging; LV, left ventricle; RV, right ventricle; PA, pulmonary artery; ASD, atrial septal defect.

## 4 Discussion

This is the first study to evaluate ventricular energetics using PV loops in a homogenous group of patients with ccTGA/IVS undergoing LV retraining for DSO, and to use a novel PV-loop derived metric of LV preparedness (LV:RV ePVA ratio) to predict outcomes of DSO. The principal finding is that patients with an LV:RV ePVA ratio ≥ 0.67 had significantly better outcomes compared to those with lower ratios. This metric is particularly valuable for patients with borderline LV:RV pressure ratios, where traditional measures may not provide clear guidance. We also show that this novel PV-loop derived metric of LV preparedness can be reliably calculated using simple cardiac catheterization and MRI derived inputs which can be obtained at all centers performing DSOs without the specialized equipment and expertise required for conductance catheter-derived PV loop recordings.

One of the challenges in determining LV preparedness for DSO is that there are no universally agreed norms with different centers using different criteria for determining LV preparedness. For example, the Stanford University program utilizes an LV:RV pressure ratio of 0.9 along with LV EDP <12 mmHg, ejection fraction >55%, indexed LV mass of >50 g/m2, and mild or less mitral insufficiency as their criteria^7,8^ whereas a pressure ratio of >0.8 has been considered adequate by the group at the German Pediatric Heart Center^13^. Among these metrics, only the LV:RV pressure ratio incorporates a patient-specific internal benchmark: the systemic RV pressure. Other metrics, such as ejection fraction, evaluate the LV in isolation, relying on ‘normal’ values for a systemic LV to judge the adequacy of a subpulmonary LV. These load-dependent metrics fail to account for the patient-specific systemic workload, limiting their ability to accurately predict the LV’s preparedness for systemic work post-DSO. The current study found that the only metrics predictive of outcomes were those that benchmarked the patient’s systemic RV—specifically, the LV:RV pressure ratio and the novel LV:RV ePVA ratio..

The ePVA ratio builds upon the pressure ratio by incorporating both the pressure and volume displaced by the ventricle, offering a comprehensive measure of total contractile energy expenditure. Systemic RV PVA measures the total contractile energy expenditure of a particular patient’s systemic circulation, and correlates closely with myocardial V̇O_2_ under a variety of loading and contractile conditions.^9,14–16^ In addition to accounting for the pressure displaced by the ventricle, the PVA also accounts for volume displaced (stroke work) and the potential energy of contraction. Therefore, comparing the subpulmonary LV PVA to the RV PVA provides a more comprehensive metric of its adequacy to perform that work after DSO than the pressure ratio alone.

There is a heterogeneous response of LVs to similarly tight PA bands, which remains incompletely understood. Some factors influencing this variation are better understood, such as atrial level shunts that introduce volume overload to the subpulmonary LV. This is a mechanism that can induce LV dilation, as illustrated in patient B (Figure 4), where the LV enlarged to become equal in volume to the systemic RV whereas typically subpulmonary LVs are smaller than systemic RVs in this patient population even with near-systemic LV pressure (such as Figure 4, patient A). However, other factors contributing to the diverse LV responses to PA banding are less well understood. For example, patient C developed suprasystemic pressure without any LV dilation. These differing responses demonstrate the complexity of LV adaptation to PA banding, where even patients undergoing similar interventions can present with a wide range of outcomes. This variability underscores the need for more precise biomarkers and metrics, such as the LV:RV ePVA ratio, to better predict LV preparedness for DSO.

## 5 Limitations

Though there was a very low overall bias for ePVA compared to mPVA, we should note that there was a positive bias at low values of stroke work and PVA. However, there are few patients who are actually referred for DSO when their stroke work and PVA are in these ranges (i.e. infants). Similarly, the Phase 1 cohort was predominantly young children aged median 2.5 years and so the equations derived from these data may not be applicable to older children.

Patients with a pacemaker in situ pre-DSO are usually unable to undergo cardiac MRI during the pre-DSO evaluation. At our center, we attempt to perform a 3D echocardiogram to assess ventricular volumes in such cases, but there may still be an underrepresentation of patients with pre-DSO pacemakers in our cohort. None of the 6 patients in our cohort with adverse outcome had pacemakers in place. This may introduce a bias because pacemaker-induced LV dysfunction is a well-described sequela of heart block in this patient population, and a distinct mode of LV failure separate from lack of LV preparedness.

In patients with severe tricuspid regurgitation pre-DSO, the systemic RV may be severely dilated. In such patients, systemic RV PVA may not be a good index for benchmarking LV preparedness as the RV PVA is falsely elevated. In such cases, one option is to calculate the PVA for an idealized ‘median LV’ with pressures and volumes at a median level for the patient’s body surface area, and then calculate the patient’s LV:LV_median_ PVA ratio. This approach has not yet been verified for outcome analysis.

Finally, though this is one of the largest homogenous cohorts of ccTGA/IVS patients reported, our sample size is still small. Multicenter validation is an important next step in understanding the generalizability of our findings.

## 6 Conclusion

The LV:RV pressure ratio remains the simplest and most reliable metric for assessing LV preparedness and should be the primary criterion. Patients with isosystemic or suprasystemic LV pressures are highly likely to be prepared for DSO. However, for patients with borderline pressure ratios (80–90%), the LV:RV ePVA ratio provides critical additional insight. While our data indicate an LV:RV ePVA ratio ≥0.67 as a marker of readiness, we recommend a more conservative clinical cutoff of ≥0.75 to minimize the risk of premature DSO, given its irreversible nature and high stakes.

## Data Availability

All data relevant to the manuscript are available upon reasonable request from the authors

## Non-standard Abbreviations and Acronyms

ccTGA: congenitally corrected transposition of the great arteries
IVS: intact ventricular septum
DSO: double switch operation
PAB: pulmonary artery band
PVA: pressure-volume area
ePVA: Estimated pressure-volume area
mPVA: Measured pressure-volume area
SW: stroke work
PE: potential energy
PSP: peak systolic pressure
ESP: end-systolic pressure
EDP: end-diastolic pressure
MAP: mean arterial pressure
SV: stroke volume
EDV: end-diastolic volume
ESV: end-systolic volume

## 7 Acknowledgments

The authors would like to acknowledge Kai-ou Tang, MA for preparing supplemental figures 1 and 2 of the manuscript. We would like to express our appreciation to all the staff at Boston Children’s Hospital who assisted in the care of these patients.

## 8 Sources of Funding

None

## 9 Disclosures

None

## Appendix

### Stroke Work, Potential Energy, and Pressure-Volume Area estimation methodology

Stroke Work (SW) is the area bounded within a PV loop (supplemental figure 1). In a normal heart, LV SW can be estimated as mean arterial pressure (MAP) * stroke volume.^11^ This relationship holds because aortic diastolic pressure is relatively high compared to systemic LV peak pressure, and thus aortic MAP becomes a good surrogate for the average afterload faced by the systemic LV during ejection. Patients with ccTGA/IVS undergoing LV preparation for DSO have LV outflow tract obstruction in the form of a PA band or native LV outflow tract obstruction. Although peak LV pressure may be high, PA diastolic pressure is very low compared to peak LV pressure and therefore the MAP in the short segment of PA proximal to the band is quite low compared to LV peak systolic pressure. Thus, the PA MAP is no longer a good surrogate of average load during ejection. Additionally, an accurate pressure recording is sometimes difficult or impossible to obtain in the short segment of main PA below the PA band. Therefore, the MAP * stroke volume approach cannot be used directly in banded subpulmonary LVs. However, since SW is the area of the PV loop, it must have some relationship with the product of LV stroke volume and LV peak pressure, with a correction factor that needs to be determined (supplemental figure 2). With this principle in mind, we derived a linear regression relationship between the product of LV peak systolic pressure and LV stroke volume, and directly measured SW. The resultant relationship is shown in supplemental figure 4 (left panels).

Potential Energy (PE) is the energy leftover in the myofilaments at the end of systole, sometimes referred to as ‘wasted work’ (supplemental figure 1). It is calculated as 0.5 * end-systolic pressure (ESP) * end-systolic volume (ESV). End-systole is defined as the upper left corner of the pressure volume loop where the ratio of pressure to volume is highest in the cycle. In a normal heart, LV ESP is approximately 0.9 * peak LV (or aortic) systolic pressure.^12^ However, this relationship assumes that there is no significant ventricular outflow tract obstruction. This relationship may not hold in the subpulmonary LV with a PAB or native LV outflow tract obstruction. Therefore, subpulmonary LV ESP was estimated (ESP_est_) as a function of LV PSP using linear regression with directly measured ESP by conductance catheter as reference. The resulting relationship is shown in supplemental figure 3 and is quite similar to the corresponding relationship previously shown to exist between the peak and end-systolic pressure in a normal systemic LV.^12^ Using ESP_est_ thus obtained, LV ePE was calculated and compared to directly measured PE (supplemental figure 4, middle panels). ePVA was then calculated as the sum of eSW and ePE, and compared to directly measured PVA calculated as the sum of directly measured SW and PE (supplemental figure 4, right panels). The regressions yielded strong and linear relationships with low bias.

**Supplemental Figure 1.**
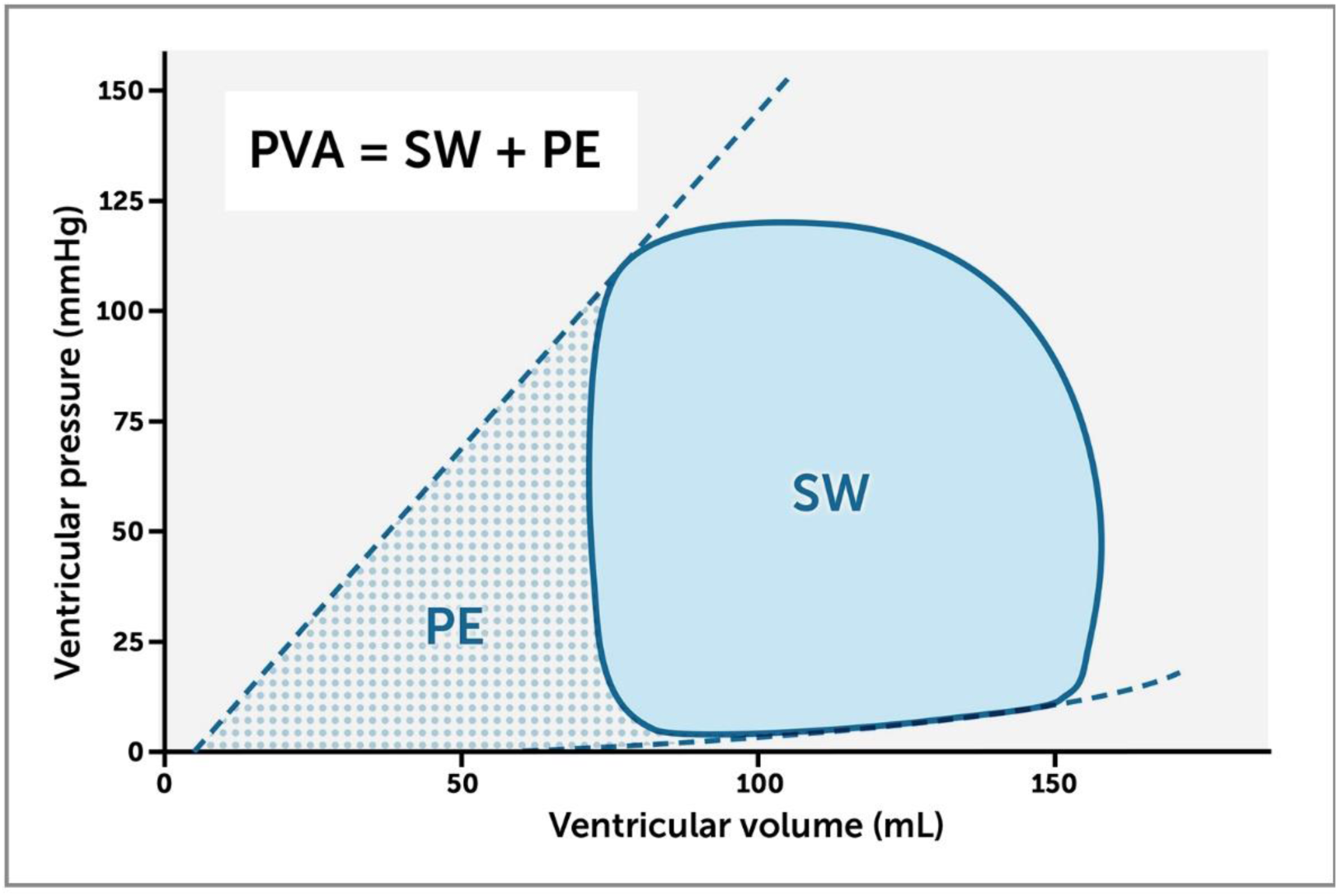
Stroke work, potential energy, and pressure-volume area (PVA). Stroke work is the area bounded within a pressure-volume loop. Potential energy is the energy leftover in the myofilaments at end-systole which is not utilized for external work and is represented by the triangular area on the origin side of the PV loop bounded by the end-systolic PV point and V_0_ (the volume-axis intercept of the end-systolic pressure volume relationship). PVA is the sum of these two components and represents the total mechanical energy of contraction.

**Supplemental figure 2.**
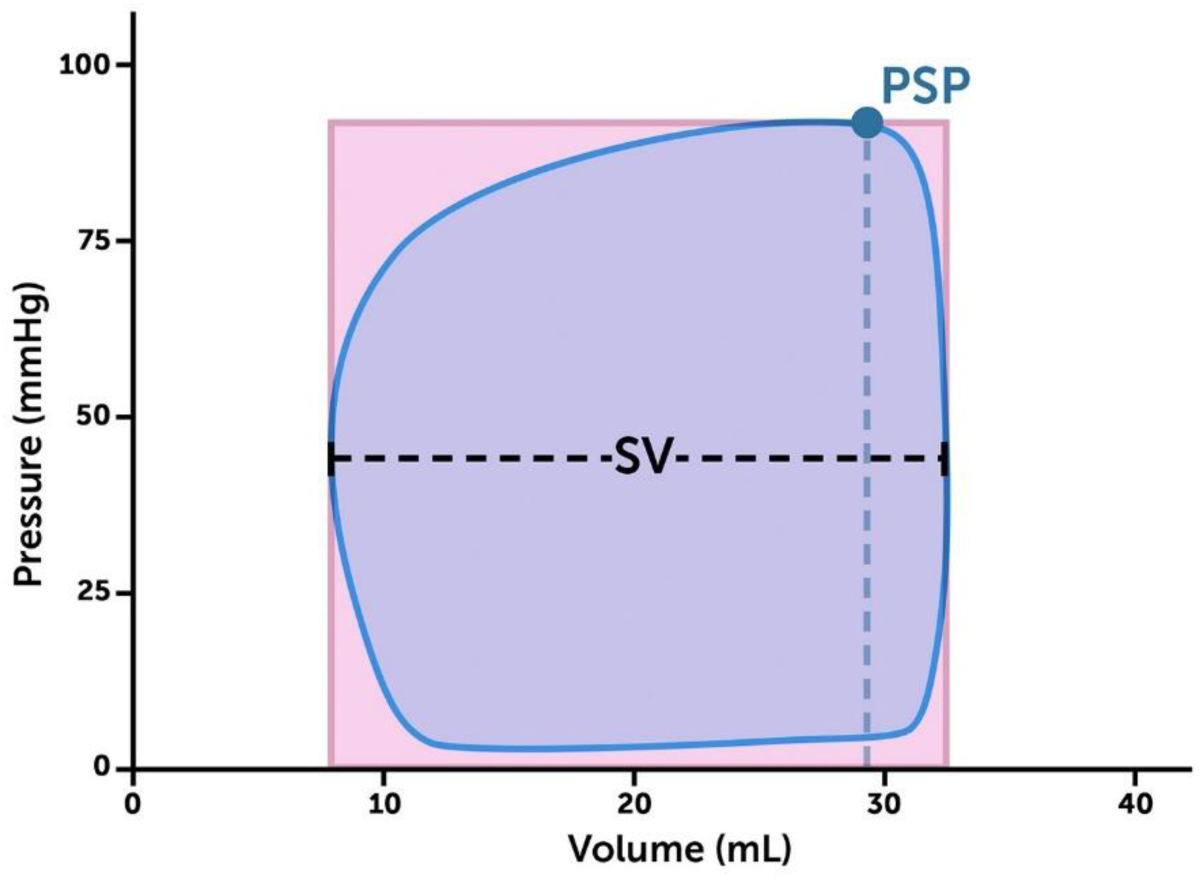
Stroke work is the area bounded within a pressure-volume loop (purple shaded area). It bears some relationship to the product of peak systolic pressure and stroke volume (pink rectangle) with a correction factor that was determined as described in the text. Abbreviations: SV, stroke volume; PSP, peak systolic pressure.

**Supplemental Figure 3.**
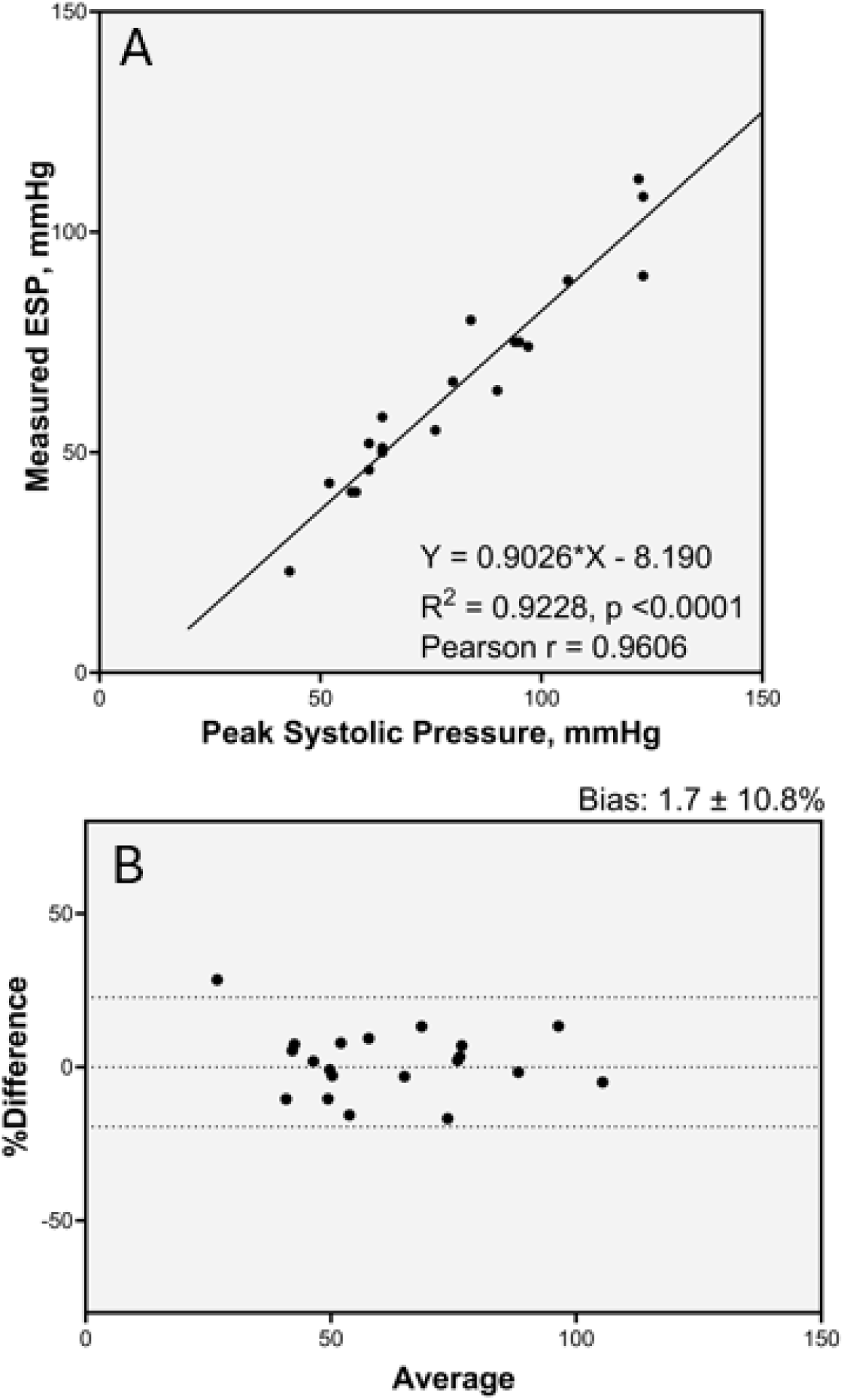
Subpulmonary LV end-systolic pressure estimation. (A) Scatterplot of relationship between directly measured LV end-systolic pressure (ESP) and LV peak systolic pressure, and (B) Bland-Altman analysis of directly measured and estimated ESP. Dotted lines represent line 95% limits of agreement in Bland-Altman analysis. Abbreviations: LV, left ventricle; ESP, end-systolic pressure

**Supplemental figure 4.**
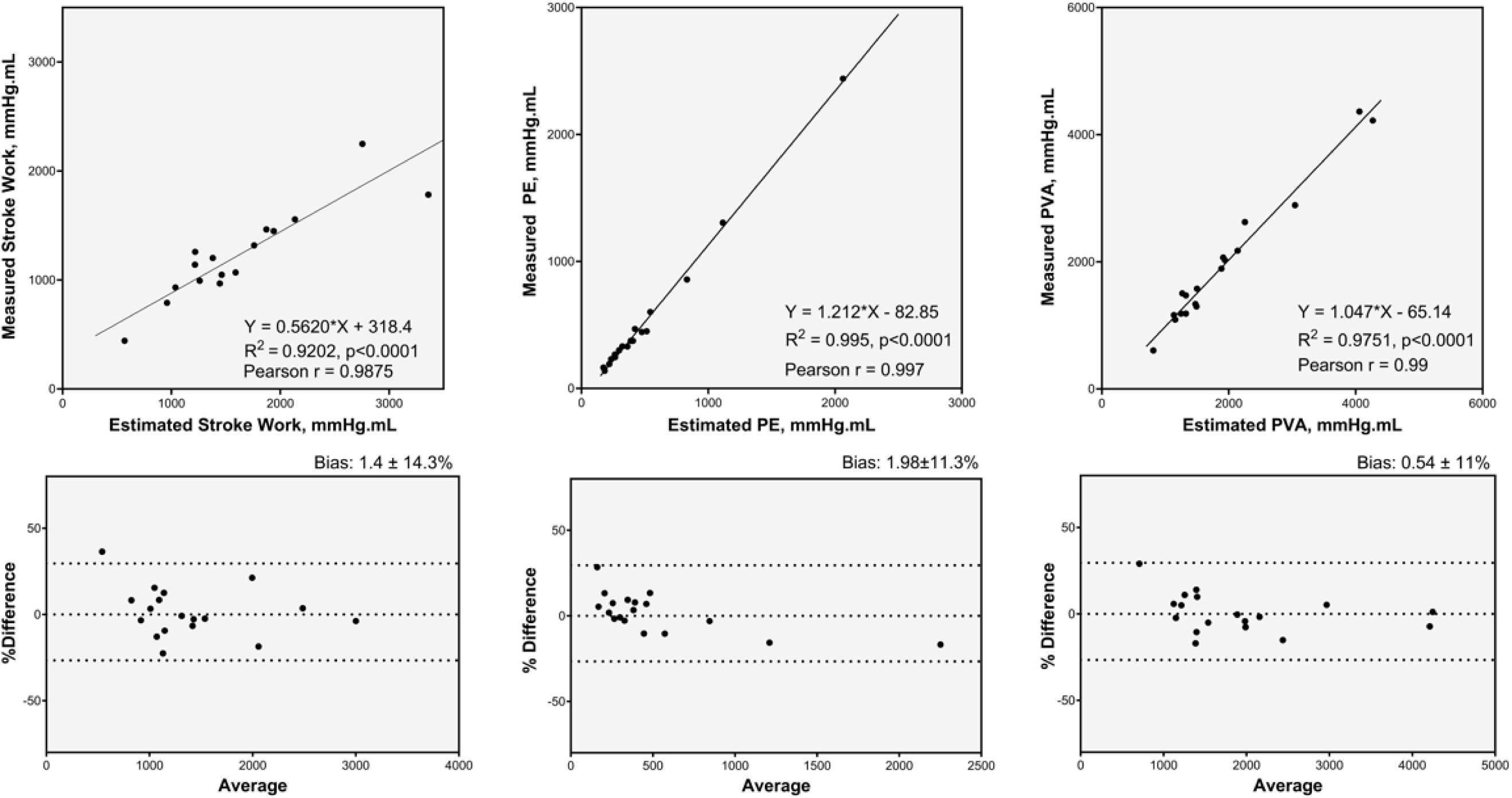
Stroke Work, Potential Energy, and Pressure-Volume Area Estimations. Scatterplots (top panels) and Bland-Altman analyses (bottom panels) of relationship between directly measured and estimated stroke work (SW), potential energy (PE), and pressure volume area (PVA). Dotted lines show 95% limits of agreement in the Bland-Altman analyses. Abbreviations: PE, potential energy; PVA, pressure-volume area

## Notes

### Competing Interest Statement

The authors have declared no competing interest.

### Funding Statement

No external funding was received in the performance of the work and the preparation of the manuscript

## References

1. van Dissel AC, Opotowsky AR, Burchill LJ, et al. End-stage heart failure in congenitally corrected transposition of the great arteries: a multicentre study. Eur Heart J. Published online August 18, 2023:1–14. doi:10.1093/eurheartj/ehad511

2. Cui H, Hage A, Piekarski BL, et al. Management of Congenitally Corrected Transposition of the Great Arteries with Intact Ventricular Septum: Anatomic Repair or Palliative Treatment? Circ Cardiovasc Interv. 2021;14(7):E010154. doi:10.1161/CIRCINTERVENTIONS.120.010154

3. Marathe SP, Chávez M, Schulz A, et al. Contemporary outcomes of the double switch operation for congenitally corrected transposition of the great arteries. In: Journal of Thoracic and Cardiovascular Surgery. Vol 164. Elsevier Inc.; 2022:1980–1990.e7. doi:10.1016/j.jtcvs.2022.01.049

4. Hraska V, Vergnat M, Zartner P, et al. Promising Outcome of Anatomic Correction of Corrected Transposition of the Great Arteries. Annals of Thoracic Surgery. 2017;104(2):650–656. doi:10.1016/j.athoracsur.2017.04.050

5. Hraska V, Woods RK. Anatomic Repair of Corrected Transposition of the Great Arteries: The Double Switch. Semin Thorac Cardiovasc Surg Pediatr Card Surg Annu. 2019;22:57–60. doi:10.1053/j.pcsu.2019.02.003

6. Mainwaring RD, Patrick WL, Arunamata A, et al. Left ventricular retraining in corrected transposition: Relationship between pressure and mass. Journal of Thoracic and Cardiovascular Surgery. 2020;159(6):2356–2366. doi:10.1016/j.jtcvs.2019.10.053

7. Ibrahimiye AN, Mainwaring RD, Patrick WL, Downey L, Yarlagadda V, Hanley FL. Left Ventricular Retraining and Double Switch in Patients With Congenitally Corrected Transposition of the Great Arteries. World J Pediatr Congenit Heart Surg. 2017;8(2):203–209. doi:10.1177/2150135116683939

8. Mainwaring RD, Patrick WL, Ibrahimiye AN, Watanabe N, Lui GK, Hanley FL. An Analysis of Left Ventricular Retraining in Patients With Dextro- and Levo-Transposition of the Great Arteries. Annals of Thoracic Surgery. 2018;105(3):823–829. doi:10.1016/j.athoracsur.2017.11.047

9. Suga H. Ventricular energetics. Physiol Rev. 1990;70(2):247–277. doi:10.1152/physrev.1990.70.2.247

10. Takaoka H, Takeuchi M, Odake M, Yokoyama M. Assessment of myocardial oxygen consumption (VO2) and systolic pressure-volume area (PVA) in human hearts. Eur Heart J. 1992;13(SUPPL. E):85–90. doi:10.1093/eurheartj/13.suppl_e.85

11. Knaapen P, Germans T, Knuuti J, et al. Myocardial energetics and efficiency: current status of the noninvasive approach. Circulation. 2007;115(7):918–927. doi:10.1161/CIRCULATIONAHA.106.660639

12. Kelly RP, Ting CT, Yang TM, et al. Effective arterial elastance as index of arterial vascular load in humans. Circulation. 1992;86(2):513–521. doi:10.1161/01.CIR.86.2.513

13. Zartner PA, Schneider MB, Asfour B, Hraška V. Enhanced left ventricular training in corrected transposition of the great arteries by increasing the preload. European Journal of Cardio-thoracic Surgery. 2016;49(6):1571–1576. doi:10.1093/ejcts/ezv416

14. Suga H. Total mechanical energy of a ventricle model and cardiac oxygen consumption. Am J Physiol. 1979;236(3). doi:10.1152/ajpheart.1979.236.3.h498

15. Suga H, Hayashi T, Suehiro S, Hisano R, Shirahata M, Ninomiya I. Equal oxygen consumption rates of isovolumic and ejecting contractions with equal systolic pressure-volume areas in canine left ventricle. Circ Res. 1981;49(5):1082–1091. doi:10.1161/01.RES.49.5.1082

16. Suga H. Cardiac energetics: from E(max) to pressure-volume area. Clin Exp Pharmacol Physiol. 2003;30(8):580–585. doi:10.1046/j.1440-1681.2003.03879.x

